## Supplementary material for "Women’s experiences collecting and accessing water in Guatemala, Honduras, Kenya, and Zimbabwe: A mixed-methods investigation": S1 Protocol

**A mixed-methods investigation of water burden: Supplement 1 Protocol**

Bethany A. Caruso^1*^, Thea Mink^1^, Madeleine Patrick^1^, Emily Ogutu^2^, Cameron Dawkins^1^, Olivia Bendit^1^, Mahnoor Fatima^1^, Ingrid Lustig^1^, Alicia Macler^1^, Jera White^1^, Alondra Zamora^1^, Alberto Emanuel Santos López^3^, Héctor Salvador Peña Ramírez^3^, Carlos Daniel Sic^3^, Jorge Lemus Chávez^4^, Sandra Antonio^5^, Jazmina Nohemí Irías^5^, Gladys Ramos^6^, Everlyne Atandi^7^, Peter Mwangi^7^, Peter Koome^8^, Rohin Otieno Onyango^8^, Petronilla Andiba Otuya^8^, Paul Ruto^8^, Morris Chidavaenzi^9^, Jammaine Jimu^9^, Sithandekile Maphosa^9^, Makaita Maworera^9^, Munyaradzi Damson^10^, Sheela S. Sinharoy^1^

^1^ Hubert Department of Global Health, Rollins School of Public Health, Emory University, Atlanta, GA, USA

^2^ Gangarosa Department of Environmental Health, Rollins School of Public Health, Emory University, Atlanta, GA, USA

^3^ World Vision Guatemala, Guatemala City, Guatemala

^4^ Centro Universitario de Occidente, de la Universidad de San Carlos de Guatemala, Quetzaltenango, Guatemala

^5^ World Vision Honduras, Tegucigalpa, Honduras

^6^ Universidad Nacional Autónoma de Honduras, Tegucigalpa, Honduras

^7^ World Vision Kenya, Nairobi, Kenya

^8^ St. Paul’s University, Nairobi, Kenya

^9^ World Vision Zimbabwe, Harare, Zimbabwe

^10^ Datalyst Africa, Harare, Zimbabwe

* Corresponding author

 (BAC)

**S1 Protocol.** Protocol for go-along interviews, semi-structured observations, and research equipment

### I. Water Journey Materials

- Participant Demographic Information Sheet
- Go-along Interview Guide
- Semi-structured Observation Guide
- Garmin 255 watches (2 per Water Journey)
- Seca 876 scales (1 per Water Journey)
- 2 measuring tapes
- 2 recorders
- Extra batteries
- Pens
- Notebook

### II. Water Journey Overview

See Garmin Watch and Scale protocols below for in-depth use of the devices (section III).

1. **Recruitment Day**
   a. Recruit participants for Water Journeys with local learning partner.
   b. Explain the Water Journey process, including the semi-structured interview, observation, and quantitative data collection with individual participants.
   c. Conduct consent process with each participant using the consent forms.
      i. Leave a copy of the consent form with the participant.
   d. Complete the Participant Demographic Sheet for each participant.
   e. Record participant height, weight, and year of birth.
      i. Enter into the Participant Watch ID spreadsheet
   f. Schedule when you will meet with the participant to conduct the Water Journey.
2. **Day before Water Journey**
   a. Sync participants’ information with the appropriate, paired participant and data collector watches (1&2) and (3&4).
   b. Prepare research devices and qualitative data collection tools; ensure that the devices are charged.
3. **Day of Water Journey**
   a. Weigh items that the participant will carry on the Water Journey.
   b. Start the Garmin Watch ‘Walk’ activity for both the participant and the data collector.
   c. Be sure to record walk start time in observation sheet, note time when the participant reaches the water source.
   d. Start the audio recorder.
   e. Conduct the semi-structured interview and observation.
      i. Depending on the circumstances, it may make most sense to conduct the interview piecemeal—instead of continuously—with some of the interview being completed before the journey, some during and some at the end.
      ii. If not continuously, be sure to:
          1. Note at what time the interview questions are being asked.
          2. Note the recorder numbers so that all files can be linked, if there are many separate interview times.
          3. Take notes along the way when interviewing is not happening so that questions can be asked related to what is being witnessed via observation. The interviewer and observer can work together to identify questions to ask about.
   f. At the water source:
      i. Record the start time for when the participant engages with water extraction/fetching related labor, which could include digging, pumping, scooping, etc. Record the time when this process is complete.
      ii. Weigh all items the participant will carry back. Make sure to label each of these items and provide notes. For example, items can include water, laundry, child (if child was weighed at start of journey, no need to re-weigh). Note how much each weigh, where / how these items are carried (on back, head).
   g. Record start time of when the participant leaves the water area to head back home. Record the time when the participant reaches their final destination.
      i. Be sure to observe any issues or challenges along the return (e.g., water spills, when and for how long there are breaks taken, if weights/items carried are shifted, moved, temporarily put down, etc.).
   h. To conclude, end the Garmin Watch ‘Walk’ activity for both watches.
   i. End the audio recorder (as applicable).
   j. Conclude and thank the participant.
4. **End of Day**
   a. Debrief with the data collection team.

#### III. Water Journey Device Details: Garmin Watches and Scales

Water Journeys will use both Garmin Watches and Scales to collect quantitative data.

#### 1. Watches: Garmin Forerunner 255

Reference the Garmin [user manual](https://www8.garmin.com/manuals/webhelp/GUID-676967A0-1B23-4384-9BC9-76F3D643F1C8/EN-US/Forerunner_255_OM_EN-US.pdf) for basic operation. Specific research protocol steps are below.

A. **Getting Started**
1. **Sign into** [**Garmin Connect**](https://connect.garmin.com/modern/)**:**

Garmin has a web-platform (website) and a phone app -- you will use both. Download the app and sign into both platforms.

Each country has 4 Garmin watches; each watch has a separate Garmin Connect account.

- All **odd** numbered watches (like G1 and G3) will be worn by **participants**
- All **even** numbered watches (like K2 and K4) will be worn by **data collectors**

Use the below watch pairings. Please **do not** mix the watches (like G1 and G4), as it will make data tracking more difficult.

- **1** (participant) & **2** (data collector) watches will be used on the **same** Water Journey
- **3** (participant) & **4** (data collector) watches will be used on the **same** Water Journey

2. **Sync** the 4 Garmin watches to your phone app by following the prompts on the app.
   a. Multiple phone accounts can be used with the same Garmin Connect account. However, only one phone can be paired with a watch at a time. This means you should select one or two point-people to sync their phone with the watches (this will be GRAs initially). You can log in and log out of the 4 watches accounts sequentially. See: [Garmin Resource](https://support.garmin.com/en-US/?faq=N7qM9eVuzl77PNYKV8ORX5#:~:text=How%20Many%20Phones%20or%20Devices,all%20the%20data%20you%20need.).

**B. Watches: Field Protocol**

**Important notes:**

- All steps with the Garmin Connect online app require **phone service** or Wi-Fi to sync with the watches. Plan accordingly by syncing participants’ information with the Garmin watches the **day before** each Water Journey when you can pair your phone.

1. **During recruitment (Day 1)**
   a. Document participant information on paper:
      i. Assign and record the **Participant Watch ID**:
         1. **Note:** This is different from the Participant ID (PID). The Participant Watch ID’s purpose is to make sure we can enter participant information that we collect during recruitment into their specific Garmin watches.
      ii. Document: Year of birth
      iii. Document: Participant weight (use scale)
      iv. Document: Participant height (use tape measure)
   b. Following recruitment, transfer the above information to the below Participant Watch ID spreadsheets:
      i. Select which recruited participant will use which participant watch (1 or 3).
      ii. This information will help retain recruited participants’ information so it can be synced with the Garmin watches.
2. **Day before Water Journey**
   a. On the Garmin Connect App (or web platform) when you have service:
      i. Make sure your phone Bluetooth is **ON** to enable syncing with watches
         1. Settings > Phone Permissions > Bluetooth is on
      ii. Log into the Garmin Connect Account for watch 1 (participant) and 2 (data collector).
      iii. Open your country’s Participant Watch ID spreadsheet (linked above in 1c) to reference participants’ information.
      iv. Input the participant’s year of birth, weight, and height into Garmin Connect.
          1. Settings > User Settings > Update height, weight, and year of birth
          2. Ensure the watch (or web-platform) has synced with the phone.
             a. The data collector watch will estimate the participant watch, and needs to have the **same** height, weight, and year of birth synced.
      v. Repeat steps iii and iv for watch 3 (participant) and 4 (data collector).
      vi. Charge all Garmin Watches overnight.
3. **Day of Water Journey**
   a. On the participant and data collector Garmin Watches:
      i. Ensure that watches 1&2 and 3&4 are used together during Water Journeys.
      ii. Ensure that GPS is **ON** for all watches.
          1. The GPS bar at the top of the watch should be green (not red).
          2. You will need to be outside for the GPS to connect.
      iii. **To start WJ:**
           1. Start a ‘Walk’ activity by holding the ‘Start’ button until ‘Walk’ shows on the screen.
           2. Press the ‘Start’ button twice to start. The watch should start recording – it will vibrate and the timer will start.
           3. Record start time and participant ID in Observation Guide.
      iv. **To end WJ:**
          1. End the ‘Walk’ activity by hitting the ‘Start/Stop’ button – the watch should vibrate.
          2. **Save the activity**: toggle the ‘Down’ button to the Save option in the menu, then click the ‘Start’ button to accept the save.
          3. Record end time and participant ID in the Observation Guide.
   b. On the Garmin Connect app:
      i. **Sync** each watch with a phone after each Water Journey ends.
         1. **This is important** to access the data on Garmin Connect!
   c. On Garmin Connect web-platform for each watch:
      i. Change the Activity Name for the **participant’s** activity to the **participant ID**
         1. **This is important** for data management!
      ii. Change the Activity Name for the data collector’s activity to **dc + participant ID**
   d. Collect Garmin watches from participants at the end of each data collection day.

**C. Watches: Exporting Data from Garmin Connect**

Each Water Journey will produce 2 files from each participant watch and 2 files from each data collector watch.

1. **Export** CSV and TCX files for participants and data collectors **each day** after data collection.
   a. On the Garmin Connect web-platform:
      i. Download the **Activity CSV file:**

(*This is a summary file of all activities.)*

1. Activities in sidebar > All Activities > Download CSV at top right corner.
2. Rename file to the standard file name convention
3. Upload file to OneDrive
4. Record that the file is uploaded to OneDrive on the data tracking sheet.
ii. Download the **TCX file**:

*(This file is more detailed than the CSV and includes GPS coordinates.)*

1. Activities in sidebar > All Activities > Click on a specific participant’s activity > Click the settings icon > Click ‘Export to TCX’
2. Rename file to standard file name convention.
3. Upload file to OneDrive
4. Record that the file is uploaded to OneDrive on the data tracking sheet

#### 2. Scales: Seca 876

#### A. Scales: Field Protocol

Reference the [scale user manual](https://www.seca.com/fileadmin/documents/manual/seca_man_876_int.pdf) to operate scale. The research protocol is below.

1. **Day before Water Journey:**
   a. Ensure that the scale can turn on and has enough battery.
   b. Each Water Journey will have 1 scale, which will be carried by the data collection team.
2. **At start of Water Journey:**
   a. Weigh what the participant will carry: empty water containers, laundry, children (if carried), etc.
      i. Use the 2 to 1 program on the scale to weigh children while the participant holds them.
   b. Record weights in observation guide to 1 decimal place.
3. **At water source:**
   a. Weigh full water containers and wet laundry.
   b. Document weights in observation guide.

**B. Scales: Data Management**

1. **At the end of each data collection day:**
   a. Input observation guide scale metrics in the Scale Data Excel sheet
   b. Record that the scale file is updated on the data tracking sheet
