## Supplementary material for "Women’s experiences collecting and accessing water in Guatemala, Honduras, Kenya, and Zimbabwe: A mixed-methods investigation": S1 Table

**A mixed-methods investigation of water burden: Supplement 1 Table**

Bethany A. Caruso^1*^, Thea Mink^1^, Madeleine Patrick^1^, Emily Ogutu^2^, Cameron Dawkins^1^, Olivia Bendit^1^, Mahnoor Fatima^1^, Ingrid Lustig^1^, Alicia Macler^1^, Jera White^1^, Alondra Zamora^1^, Alberto Emanuel Santos López^3^, Héctor Salvador Peña Ramírez^3^, Carlos Daniel Sic^3^, Jorge Lemus Chávez^4^, Sandra Antonio^5^, Jazmina Nohemí Irías^5^, Gladys Ramos^6^, Everlyne Atandi^7^, Peter Mwangi^7^, Peter Koome^8^, Rohin Otieno Onyango^8^, Petronilla Andiba Otuya^8^, Paul Ruto^8^, Morris Chidavaenzi^9^, Jammaine Jimu^9^, Sithandekile Maphosa^9^, Makaita Maworera^9^, Munyaradzi Damson^10^, Sheela S. Sinharoy^1^

^1^ Hubert Department of Global Health, Rollins School of Public Health, Emory University, Atlanta, GA, USA

^2^ Gangarosa Department of Environmental Health, Rollins School of Public Health, Emory University, Atlanta, GA, USA

^3^ World Vision Guatemala, Guatemala City, Guatemala

^4^ Centro Universitario de Occidente, de la Universidad de San Carlos de Guatemala, Quetzaltenango, Guatemala

^5^ World Vision Honduras, Tegucigalpa, Honduras

^6^ Universidad Nacional Autónoma de Honduras, Tegucigalpa, Honduras

^7^ World Vision Kenya, Nairobi, Kenya

^8^ St. Paul’s University, Nairobi, Kenya

^9^ World Vision Zimbabwe, Harare, Zimbabwe

^10^ Datalyst Africa, Harare, Zimbabwe

* Corresponding author

 (BAC)

**S1 Table.** Thematic descriptions of women’s water collection experiences and practices, by community

| **S1a Table. Guatemala** | | | | | | | |
| --- | --- | --- | --- | --- | --- | --- | --- |
| **Community** | **1 n=3** | **2 n=4** | **3 n=3** | **4 n=3** | **5 n=3** | **6 n=3** | **7 n=3** |
| **Water Sources Visited During Data Collection** | Water sources visited included a river and a well. | Water sources visited included a river and a well. | Water sources visited included a spring, a river, and a well. | Water sources visited included a spring and a tank. | All participants visited a public tank in the center of the nearby municipality, where they must pay for water. | Water sources visited included a public tank/pila and a spring. | All participants visited a public tank. |
| **Other Water Sources Used by Participants, But Not During Data Collection** | Additional water sources mentioned include a river near the community, a more distant river if needed because of crowds or reduced supply in the dry season, well water, a spring that typically dries up in the summer, and rainwater collection when the weather permits it. | Additional water sources mentioned include rainwater harvesting into a large tank, river, a secondary river during dry season that is farther away and requires a minibus, and a well owned by a neighbor. | Additional water sources mentioned include a spring where a pipe was installed for access, rainwater collection when the weather permits, a source at a family member's home, a river, a well, and a ravine. | Additional water sources mentioned include a recent connection to water at the participant's house, well water, rainwater collection, rainwater collection, paid water from a nearby community source, and a source at a family member's house where water is stored in a large plastic tank, brought there by a type of mechanical well. | Additional water sources mentioned include rainwater collected through pipes into a household pila, piped water that comes twice a week or for half an hour in the afternoon or in small amounts every two days, well water, spring water, a tanker which come once a month to provide water, and a community well. | Additional water sources mentioned include a pond built by the community, water sources at a family member's house, rainwater collection, a community tank where water must be paid for, depressions in the ground where covered spring water pools to create a small pond, and pilas belonging to a neighbor. A participant mentioned that they had formerly had pipes connecting a spring to their houses, but after a construction project, they were no longer allowed to connect their pipes to the spring, though the owner still allows them to wash at the spring for free. | Other water sources mentioned included a water tank, water provided by neighbors, rainwater collection, a tap in the yard or household water connection, a community-owned system requiring payment, a spring beyond a ravine, and a pond which dries up in the summer. |
| **Method of collection** | In this community, women were mostly observed going to the source where they do laundry. One observation was at a well, where the participant uses a bucket tied to a rope to get water. | Water collection (instead of washing) was observed for two participants. One looked for the cleanest spot in the river and then filled her containers there. Another pulled water from the well, which she reported requires strength and is easier with her daughter to help. | One participant was observed washing clothes. For the participants collecting water from the spring and the well, water passed through a tube or pipe and was collected from the tube. One participant crouched to collect water from the tube. | The method of water collection was not documented. | The method of water collection was not documented. | The method of water collection was not documented. | The method of water collection was not documented. |
| **Number and timing of trips** | Participants reported collecting water every day in 1-12 total trips. One participant reported that she does 4 trips, and her husband helps with another 7 or 8. The timing of trips depends on household chores, weather, and the children's school schedules. Fewer trips are required in the winter when rainwater is also collected. | Participants reported collecting water from 2 to 5 times a day in the morning or afternoon, or as many as 10 times a day if water is needed for bathing. If women bathe in the river, they may take additional trips to the water source but not collect additional water. Sufficient drinking water may be collected in 1 or 2 trips a day. The frequency of water collection decreases if enough rainwater has been collected. | Participants report collecting water between once a day from the well and 6 times a day from the spring, but less in the rainy season because they collect rainwater. They collected water as needed, in the morning or afternoon depending on when they have enough and when it's raining. | The number of times that participants reported collecting water varied from twice a week in the morning, to once a day after lunch, to eight times a day beginning at 5:30 in the morning. One participant collects water infrequently because she has to pay for transportation to and from the water source. | Two participants reported that the water for collection is only available twice a week for 30-45 minutes, so they have to collect water at those times. Two participants reported that the water for laundry at the tank runs out, so they go early in the morning to do their laundry. One participant reported that she collects water 3 times a day in the afternoon but fetches less when it is raining. | Participants made separate journeys to sources to wash or collect water. They reported as few as 3 and as many as 30 trips to the water source to do laundry and collect water, depending on season and what the household needs are. Participants reported needing to take more trips if the family was bathing, or if they were collecting water to fill their pila at home. One participant reported that sometimes they get to the water source to do laundry, and it is so crowded they do not have space to wash, so they have to return home without washing their clothes. | Participants reported making between 3 and 6 times a day, with up to 9 trips in the summer. They may fetch water in the morning or after lunch. |
| **Method of carriage** | Women carried pots of water. One participant reported that her husband sometimes helped when they all needed baths. One transported clothes in sheets. A participant who was pregnant was followed twice, once for laundry and once for water collection; she carried laundry on her back and carried water in a pot on her head. She noted that she carries less water (only one pot), and her husband now also makes many trips to collect water because of her pregnancy. | One participant carried laundry home in a bag and carried water pots with ropes or balanced on their heads. Water was collected in pots and barrels. | Women carried medium or large pots. Participants reported that children may also fetch water in smaller pots if they are present. Laundry was wrapped in a sheet. | All participants used big pots or buckets. One participant carried two up to the source but carried them back one by one. One participant carries water on her head and her daughter in arms, necessitating breaks. One participant reported that either she pays for transportation, or her husband borrowed a car to collect water together. | Two women carried water on their heads; one of them used a cloth to hold the pot on her head. Another held her pots by their handles. One participant mentioned she purchases water by the barrel but then pours it into pots to transport it home because the barrel is too large to transport without a car. One reported transporting a barrel in a vehicle. Children carried small pots. | Participants carried the water pots on their head cushioned with a "yagual" or rolled up blanket. One participant reported that she occasionally hires a transportation service to take back water and clothes, or her brother takes her in his car. She said that sometimes she has to leave pots behind to be collected later. Another reported that sometimes her husband or daughter helps her carry clothes back home because they are too heavy to handle by herself now that she is pregnant. | Participants carried water pots on their head; one participant reported that when she has two pots, she carries one on her head and one with her arms. |
| **Terrain and risks** | A water source was far away, and the terrain to get there was mountainous, with a slippery and unpaved road. Participants reported that when it rains, the river overflows and is dangerous. Two participants reported having slipped and fallen in the past; one participant noted that she and her daughter both frequently fall, and once she broke a full water pot. There are no houses near the water source, and there have been suspicious people and snakes. One participant reported that a man was murdered at the river recently, so she now always brings someone with her. | Participants reported using less steep and bumpy paths when they had options. There are no safety issues like animal attacks, but older children go with younger children. It is slippery, and there are many holes in the path. There are snakes. The path is owned by other people, and they cannot improve the path without permission in case the owners close the path. One participant reported that her daughter slipped and fell while carrying a large pot and broke her leg when her foot was trapped. | Participants reported that the ravine was closer than the well, but the path was more challenging. The terrain was steep and slippery, and it is dangerous when it rains because the current is strong. Participants expressed fear that they or children could fall into water sources or ravines. There are snakes; one participant reported that her daughter had been bitten by a snake. | The path was steep and going uphill was difficult. Participants reported that the path is slippery and dangerous in the rain. One participant reported that wolves can appear at night, but she had never encountered one. | Participants reported possibilities of dangerous people and barking dogs. | The path to the water sources became slippery and muddy in the rain. Participants reported landslides in the rainy season. Sloped areas required participants to walk slowly to avoid falling. One participant said that the path became more difficult when corn in the field has grown. Participants noted that children can have accidents, and that there is a risk of attack from animals and people. | Paths were steep and slippery, with an unpaved downhill section that is covered in mud and particularly challenging, especially in the rain. Participants reported that there is a risk of slipping and falling, and that tree branches break and fall when it is windy. One participant fell while carrying the water container and sprained or dislocated her ankle. Participants reported the presence of dogs in the area that can come out and bite. |
| **Physical effects of water collection** | Participants reported discomfort when walking and carrying heavy things because of low blood pressure and pain in feet at night. One participant was 6 months pregnant and reported experiencing more tiredness and pain as a result and needing her husband to help carry laundry. | One participant reported that her daughter slipped and fell while carrying a large pot and broke her leg when her foot was trapped. | Participants reported backache, pain in hands and feet from standing in the water while washing clothes; pain in the body; and weakness due to age, diabetes, and gallbladder surgery. | One participant reported that she cannot carry heavy loads after two caesarian sections. | No data collected | Due to pregnancy, one participant had to carry a smaller pot of water and required her husband's help to carry clothes. She reported that pregnancy makes water collection more difficult and causes pain. One participant complained of body pain, soreness, and difficulty in walking long distances. | Participants reported fatigue as a result of water collection. One participant had fallen and sprained or dislocated her ankle because of a fall during slippery conditions. |
| **Children** | All participants had a child with them. One participant brought a 4-year-old who walked. One participant carried a 2-year-old on her back. The participant who was pregnant also brought her daughter with her and held her hand. When her daughter is tired, she carries her on top of the laundry. She noted that she would continue to do laundry when the pregnancy was very advanced but would look for someone to help her after the child was born. One participant reported fetching water without children during the day, going again with children in the afternoon. | Two of four participants carried a child. One participant brought a pre-teen child along. One participant reported that older children may fetch water or go with participant. One participant reported that she often sends her daughters to collect water. Participants carry buckets and children. | Two participants carried a child. One participant regularly brings children or grandchildren with her, who help by carrying small pots | All participants carried children. One participant stated that it is difficult to carry her daughter in one arm and water in the other. Participants report that older children may come and help. | Two participants brought children with them. One of them brought a three-year old and reported that her children are too young to help. The other reported that her other children occasionally come with her and carry water in small containers. Another participant did not have any children. | The participant who was pregnant also carried a child who she breastfeeds. She reported that she and her eldest daughter take turns collecting water, and when she gives birth, her daughter will take on the responsibility of collecting water. Another reported that her older child may carry a pot. | Two participants carried small children and reported that their older children sometimes help. One participant carried her daughter, but she leaves her son at home because she is afraid, he will fall in the well. Another participant's children are grown; her daughters or grandchildren occasionally help. |
| **Other work done during water collection** | Participants collected firewood and washed clothes, shoes, ponchos, and blankets. | Participants washed clothes and utensils in the river washstand or kneeled in the river. One participant reported bringing ducks to swim in the river. Clothes were carried back wet so that animals do not step on them while they dry. | Participants washed clothes at the river or ravine and carried them back wet. | Participants washed clothes and dishes. | Participants washed clothes and utensils, but this work was not during water collection. Washing and water collection were accomplished during separate trips. | Participants washed their hair and washed clothes and dishes. Clothes were carried back wet. | Participants washed clothes. One participant mentioned that she is sometimes paid to wash clothes for others. They carried clothes back wet. |
| **Perceived water quality and water treatment** | One participant said that she is careful to only use clean water because her son became ill once and the doctor warned against contaminated water, but she did not specify how she ensures her water is clean. Another participant reported that spring water is cleaner than well water because the spring is covered, whereas dogs drink from the well, but the spring dries up in the summer. Another reported that a dog had fallen in the well and contaminated it, so they were currently not using the well, but normally, the well water is cleaner, particularly in the summer, when it is emptied out and cleaned once a month using a brush and detergent. | Two participants reported that the nearer river is cleaner than the more distant river because the community nearby releases waste and sewage into that river. Well water was perceived as clean because it comes from beneath the ground, is cement-lined, and the well is cleaned every month or two. Participants reported boiling river water and rainwater in case it is dirty from the roof. They also reported sometimes boiling well water in case it gets contaminated by sewer discharge. | When collecting spring water, drinking water was collected from a higher part of the spring, where the water is freshest. The well water was perceived to be clean, as was the river water when it is not rainy season. The participant that used a well reported that her family cleans the well every Sunday. A participant that used the spring reports that they pay someone to clean the area once a month. Participants did not note steps taken to treat water after collection. | One participant said that the water she uses is clean. Participants that used a spring noted that they try to keep the area clean by digging with a hoe to allow for more accumulation of cleaner water and clearing the water of worms and debris. One participant mentioned filtering water when it is muddy from rain or landslides and putting a lid on stored water to protect water from contamination. | Users of the public tank wash it periodically when the tank is empty. One participant reported that rainwater from roofs is dirty, so it is not used for drinking or cooking. One participant reported using a filter to purify water for drinking and/or cooking. | Users of the community tanks and pilas reported collaborating around cleaning the tanks. The women or families who use it take turns. The participant who used spring water on a neighbor's property reported that users of that source take care of the pond. | Participants reported that users of the tank take turns cleaning it. There are pond managers who clean it biweekly in turns. The river water is cleaner after it has rained or when the water pipe above has been cleaned. Because the river water is dirty, it is used only for washing. Rain brings mud, brush, and garbage into the river. |

| **S1b Table. Honduras** | | | | | | |
| --- | --- | --- | --- | --- | --- | --- |
| **Community** | **1 n=2** | **2 n=3** | **3 n=4** | **4 n=3** | **5 n=3** | **6 n=2** |
| **Water Sources Visited During Data Collection** | Participants collected from a river and a spring. | Participants collected from a river, stream, and faucet. | Participants collected from a well and a spring. | Participants collected from a stream and from pooled river water. | Participants collected from a public faucet and a river. | Participants collected from a stream and a well. |
| **Other Water Sources Used by Participants, But Not During Data Collection** | Other sources mentioned included a tap, a river, a well, and rainwater collection. | Other sources mentioned included a stream, a river, and a tap at a family member's house. | Other sources mentioned included wells, streams, a source at a family member's house, and rainwater harvesting. | Other sources mentioned included a sink, a river, and a stream that is dry in the summer but has water in the rainy season. | Other sources mentioned included a public faucet/tap, a river, a stream, purchased sachets, neighbors that provide drinking water, paying for others to collect water, and rainwater collection. | Other sources mentioned include a stream, a well, and purchased water. |
| **Method of collection** | The method of collection was undocumented | The method of collection was undocumented. In winter, a participant reported getting water through a hose. | Women collected water from a pipe with a strainer at the end and a hose connected to a spring. | One participant collected water from the stream using a bucket; for all other participants the method of collection was not documented. | The method of collection was undocumented. | The participant used a manual pump. |
| **Number and timing of trips** | Participants reported collecting water 2-4 times a day. There is a water system that receives water every day in the winter but often fails in the summer. | Participants reported collecting water more often in summer dry months. One participant collected twice every day, in the morning and the afternoon. Another collected twice a week, at 5 AM to carry water to drink, and at 11 AM to wash. A third collected 6 times a day and reported going to a source at a family member's house every 3 days. | Participants reported fetching more often in summer dry months. Three of the participants collect water 2-3 times a day, depending on how much water is needed. One participant only collects every 3-4 days. Participants reported choosing the timing of their water collection to avoid mosquitoes or the rain. | Participants reported fetching more often in the summer months. One participant reported only collecting once or twice a week but filling many containers. Other participants reported going 3-10 times a day to wash and collect water. One participant reported preferring to collect in the morning to avoid the sun. | Timing of water collection for the two participants collecting from a public tap varied based on when water is available in the tap, which is inconsistent and based on the decisions of the person in charge of the taps. Another participant reported making 5 trips a day to collect water. She reported collecting water less frequently in the dry summer months, and she avoids collecting when the sun is intense. | Both participants reported avoiding collecting water when it is sunny. One participant makes 3 trips a day to collect water from the stream and 3 trips a week to collect water from the well. The other participant reported that she goes to purchase drinking water 3 times a day, but that she only collects water from this source once every two weeks, because her household is small, and water lasts a long time. |
| **Method of carriage** | Participants carried water in a bucket. No details were documented. | Participants carried water in buckets on their heads or by handles. | Participants carried buckets with their handles. | Participants carried water in buckets on their heads or by handles. | Participants carried buckets by their handles. | Participants carried water in buckets on their heads or by handles. |
| **Terrain and risks** | Part of the journey was on a main street. Participants reported that the road becomes slippery when it rains and there are problems with falls. | The terrain was not too difficult, but the road was irregular and there were rocks along the way. Participants mentioned that there are snakes. Participants reported being careful around the water source, which rises in the rain, and making sure that children are safe around the water source. | The trail to the water source was difficult. The path was reported to be slippery when rainy, with falling branches in the wind. Participants reported that animals and snakes may appear, that there are a lot of mosquitoes, and that the place is not suitable for children or safe at night. | Participants did not report safety concerns, but they did report that the path goes up and down hill and is slippery, with excess mud in the rainy season which may lead to slipping and falling. Carrying water in those conditions is challenging. One participant reported falling in the past. | Participants reported that the ground is slippery but otherwise had no safety concerns or complaints about the terrain. | Participants reported that the path is slippery, especially when it rains, but otherwise had no safety concerns or complaints about the terrain. |
| **Physical effects of water collection** | Participants reported physical discomfort and difficulty walking long distances. | Participants reported back pain, neck pain, and exhaustion. | Participants reported fatigue, exhaustion, and leg pain. | Participants reported exhaustion, pain in the back, knees, and legs, shortness of breath, and chest problems. | Participants reported exhaustion, back problems, pain in the tailbone, and headaches. | Participants reported pain in their arms, and one example of a sprained ankle. |
| **Children** | One participant reported that her children go with her, however they were not with her on this journey. The other did not have children with her on this journey but reported that her children accompany her when she goes to a water source to wash dishes or do laundry. | All participants reported occasionally having children with them when they go to a water source. One participant reported being accompanied by children when bathing or washing clothes but not for water collection. One participant had two children with her, the older of whom carried the younger. Another participant reported that she usually collects with children, but she was alone for this journey. | Two participants reported that children go to the water source, while the third reported that only adults go to the water source. It was not documented if children were carried or accompanied by mothers on their journey. | All participants reported at least sometimes going with their children to the water source. Children may carry water. | One participant reported bringing children and two reported that they do not bring children. | One participant reported going with her child, though she did not have a child present on this journey; another reported always leaving her child with a neighbor. |
| **Other work done during water collection** | Participants reported washing clothes and utensils, bathing, bathing children, and collecting firewood. | Participants reported washing clothes and dishes, bathing children, and collecting firewood. | Participants reported washing clothes and bathing. | Participants reported washing clothes, bathing, bathing children, collecting firewood, and picking fruit. | Participants reported washing clothes, collecting firewood and picking fruit. | Participants reported washing clothes and stopping at the grocery store. |
| **Perceived water quality and water treatment** | One participant reported that the river water is contaminated by animals upstream, so she collects water as far upstream as possible; another said that the river water is clean but gets dirty when it rains. | One participant reported that the river water is cleaner in the morning; another said that it is clean in the winter, but it looks dirty when it rains | Participants reported that water from the well is clean and safe for consumption, but water in the spring is contaminated. The upper part of the stream has safer water because there is less contamination. | Participants reported that the river water gets contaminated and dirty, especially when it rains. | Participants reported that the water is contaminated. One participant used separate parts of the river for washing and collecting drinking water to avoid contamination of drinking water. | No data were collected. |

| **S1c Table. Kenya** | | | | | | |
| --- | --- | --- | --- | --- | --- | --- |
| **Community** | **1 n=3** | **2 n=4** | **3 n=3** | **4 n=4** | **5 n=3** | **6 n=4** |
| **Water Sources Visited During Data Collection** | All participants collected water from an unprotected dug well in a river. | All participants collected water from an unprotected dug well in a river. | All participants collected water from an unprotected dug well in a river. | Participants collected water from an unprotected dug well in a river or a newly constructed borehole. | Participants collected water from an unprotected dug well in a river or a newly constructed borehole. | Participants collected water from an unprotected dug well in a river or a newly constructed borehole. |
| **Other Water Sources Used by Participants, But Not During Data Collection** | Participants reported that in the rainy season, they also collect rainwater and collect water from unprotected springs. There is also a cemented well at the top of a mountain or hill that has water even in the dry season, and where rainwater collects in rocks in the rainy season. | Women reported that in the rainy season, they participate in rainwater collection and surface water collection from small dams and gullies. | Women reported that in the rainy season, they also collect surface water from small dams. | Women reported that in the rainy season of about one month, they may collect water where it accumulates in dug water pads near the home. At the top of a mountain/hill, water collects in rocks and there is a cemented well, which usually has water even in the dry season. One woman noted that she often collected water from the newly constructed borehole, though she did not visit in on the day of data collection. | Women reported that in the rainy season, they collect surface water from small streams or nearby water pots which may hold water for days. | Women reported that in the rainy season, they collect surface water from unprotected dams and streams. |
| **Method of collection** | Women dug the well at river, scooping out and throwing away the early water, which was brown and dirty, and then they collected the clear water after the well re-filled. They reported using water pans to collect water in the rainy season. | Women dug the well at river, scooping out and throwing away the early water, which was brown and dirty, and then they collected the clear water after the well re-filled. They reported using water pans to collect water in the rainy season. | Women dug the well at river, scooping out and throwing away the early water, which was brown and dirty, and then they collected the clear water after the well re-filled. | Women collecting from the river dug the well, scooping out and throwing away the early water, which was brown and dirty, and then they collected the clear water after the well re-filled. The method of collecting from the borehole was not documented. | Women collecting from the river dug the well, scooping out and throwing away the early water, which was brown and dirty, and then they collected the clear water after the well re-filled. The method of collecting from the borehole was not documented. | Women collecting from the river dug the well, scooping out and throwing away the early water, which was brown and dirty, and then they collected the clear water after the well re-filled. Three people got inside the well to fetch the water out. The method of collecting from the borehole was not documented. |
| **Number and timing of trips** | Women reported collecting water in the morning because the sun is too hot later. They mostly reported fetching once, but sometimes twice in a day when fetching from the river. | Women reported collecting water in the morning because the sun is too hot later. They mostly reported collecting once or twice a day, but in the rainy season, they reported they may go more often to smaller water sources nearby. | Women reported that they collect water once a day because of the long distance, and that they go in the morning to avoid the heat. | Women reported that when they collected from the river, they would go once or maybe twice if they had many chores requiring water. In the dry season, they usually only went once, in the early morning. In the rainy season, they reported visiting dams nearby more often, as often as 5 times a day. When using the newly constructed borehole, they reported going as often as 4 times a day. | Women reported collecting water from the river only once a day because the journey is long and may take even longer if they are bringing livestock with them. They report that they go in the morning because it is less hot, and because if they go in the afternoon, it may be dark by the time they are returning. When they collected from the borehole, they may make 2 trips a day but are limited because the borehole is only open for limited hours (from 9 or 10 A.M. until 1:00 P.M.). During the rainy season, they report visiting water pods 3 times in a day. | Women reported going to collect water from the river or mountain only once a day in the morning, when it is not as hot and they have more energy. If they collected from the borehole, they may make two trips in a day, but they are limited because the borehole is not open every day, is open for limited hours, and there are sometimes long lines. |
| **Method of carriage** | Women carried jerricans (10 L or 20L) on backs, by dragging or rolling, or using both methods at once. They reported that children may assist in rolling, and that they used to use donkeys to help collect water but most of them have died. | Women carried jerricans (10 L or 20L) on backs, by dragging or rolling using a rope, or using both methods at once. The small can is carried and the larger can is rolled. They noted that they use the 10 L more often than the 20 L because the 20 L is too heavy to move over the long distance. They reported that children may assist with rolling, and that they used to use donkeys to help carry water, but most of them have died. | Women carried jerricans (10 L or 20L) on backs, by dragging or rolling, or using both methods at once. The 10 L jerrican is carried and the 20 L jerrican is rolled. Participants noted that on rest days, just the small jerrican may be brought, but usually both containers are used. | Women carried jerricans (5 L, 10 L, or 20L) on backs, by dragging or rolling using a rope, or using both methods at once. The smaller jerrican may be tied on top of the larger jerrican on the back, particularly in areas where they can't be rolled because of the rocks and gullies. Participants noted that on rest days, just the small jerrican may be brought, but usually both containers are used. They reported that they may use the 10 L jerrican if they have leftover water from the day before or if they are on their second trip of the day. Decisions on size of jerrican also depended on level of exhaustion. They reported that donkeys used to help carry water, but they have died. | Women carried water in jerricans. When collecting at the river, they reported carrying less water, usually a 20 L or a 10 L and a 5 L at the same time.. Jerricans were tied horizontally to balance the weight and carried supported by the back and the head. | Women tied 10 L or 20 L jerricans on the back to carry them. The 10 L was often selected because of the distance, or because a baby was also being carried. |
| **Terrain and risks** | The terrain was flat, with some gullies. Carrying heavy jerricans may cause falls. There was a risk from wild animals, specifically elephants and leopards. They noted that elephants may be at the river or chase participants. | The terrain was flat, with some gullies. Participants reported that carrying heavy jerricans may cause falls. Participants noted the presence of elephants, but disagreed on if they were likely to attack. They reported that there are sometimes rumors of attacks from people, but that this is rare. | The terrain was flat and plain, with some gullies. Participants noted that the terrain may occasionally lead to falls. Participants noted that there are animals such as elephants on the way to the river, and that there are rumors of people who attack others, but no participants had personal experience of being attacked. | The terrain on the path to one river was hilly, and it was flat with small gullies on the way to another river they used. Sometimes, women fall while carrying water. Participants noted that for one source, they must climb a mountain and cross large gullies on a path that donkeys have difficulty navigating, and people may fall because of mistakes or rocks. Participants reported that wild animals such as elephants may be present and attack; on one water journey, an elephant was spotted that day. | There were no rocks or hills, but women climbed up and down deep gullies and passed through bushy areas. Participants reported that the path had risks, including wild animals, particularly elephants, and rumors of a man who attacks people, though none of them had ever seen him. | The terrain had rocks and deep gullies, which were noted to cause falls. Women climbed hills and went over sandy areas. Women reported that there were sometimes elephants, and that when jerricans are left at the borehole, they may be destroyed by donkeys or stolen. |
| **Physical effects of water collection** | As a result of water collection, women reported: sweating a lot; stomach pain; chest aches; weakened bodies; exhaustion; pain all over the body, particularly the head and neck. Water collection was noted to be particularly difficult when pregnant. | As a result of water collection, women reported: blisters on their hands from the rope; back pain; headache; sharp pains across the stomach; possible miscarriages; chest aches; and pain throughout the body, particularly in the shoulders. | As a result of water collection, women reported: fatigue; exhaustion; and pain throughout body, particularly the shoulders and legs. | As a result of water collection, women reported: exhaustion; joint pain; high heart rate; and pain in various body parts including the back, ribs, chest, shoulders, and the head, particularly where the rope presses. | As a result of water collection, women reported: back pain; exhaustion; body aches; and poor sleep. | As a result of water collection, women reported: exhaustion; pain all over the body; and headaches. One woman noted that with the new borehole, they now get more sleep and have more energy. |
| **Children** | One participant hurried on her journey because she had left her baby at home. Women reported that children may roll jerricans to collect water, particularly during school holidays and weekends. When children collect water, they go in groups. | Women reported that mothers with small children carry them while collecting water. On the way back from the source, they carry water on their backs and babies on their fronts. Children may collect water, but they do not go alone because of the risk from elephants. Women noted that young children may be left at home with older children. Children are more likely to fetch during rainy season, when there is access to water in nearer sources. | Women reported that children may help with water collection on Saturdays so the mother can rest. Mothers with small children report trying to find someone to take care of small children while they are collecting water, or having older siblings take care of younger siblings. | Women reported that small children are usually left at home with another woman or are carried during water collection. Older children may help when schools are closed on weekends. Children do not collect water alone, but with their mother or other women. Children typically go to closer locations when they collect water, and not to the more distant sources. | One participant carried a child while collecting water. Women reported that children may be left with an older child or mother-in-law, or they may accompany the mother to collect water. If they go to the water collection point, they may help carry a 5 L jerrican. | Women reported that children are usually left at home with another woman (co-wife or grandmother). They may go alone to the borehole, but not to the river because of the distance and the wild animals. |
| **Other work done during water collection** | Additional work reported or observed during water collection included: bathing children; washing clothes (which required carrying laundry to the water point); and collecting and carrying firewood. | Additional work reported or observed during water collection included: bathing children; washing clothes (which required carrying laundry to the water point); collecting and carrying firewood; and watering goats. | Additional work reported or observed during water collection included: bathing; washing clothes (which required carrying laundry to the water point); collecting and carrying firewood; watering donkeys; and cleaning jerricans. | Additional work reported or observed water collection included: bathing; washing clothes (which required carrying laundry to the water point); collecting and carrying firewood; cleaning jerricans using sand; and collecting wild fruits if visiting the mountain source. | Additional work reported or observed during water collection included: bathing children; washing clothes (which required carrying laundry to the water point); watering donkeys; and collecting and carrying firewood. | Additional work reported or observed during water collection included: bathing children; washing clothes (which required carrying laundry to the water point); watering donkeys; and collecting and carrying firewood. Participants reported that when they collect at the borehole, they are able to bring home sufficient quantities of water that they may wash clothes at their homes instead of at the water source. |
| **Perceived water quality and water treatment** | Participants reported that the water was good, was tasteless, had no odor or color, and was salty in the dry season. They attempted to collect clean water by pouring out unclean water from the well and waiting for clear water to come to the surface. Two participants reported that the soil treats the water with salt or natural chlorine. One reported sometimes boiling water if it is brown, and sometimes being provided with WaterGuard to treat the water. | Women reported that water from the dug well at the river is clean and fresh after the initial brown water is poured out. The water was reported to be soft in the rainy season but salty or hard in the dry season. Participants did not report treating water from the river, but they did treat water from the dams, which is dirty, brown, and frequently used by animals, with chlorine or WaterGuard. | Women reported that the water from the dug wells at the river is fresh and colorless when it rains, but salty and brown in the dry season. When they collected from dug wells the river, they poured out the brown water and only collected the clean water. They did not report typically treating water; one participant reported boiling water only when her children had diarrhea. | The water from dug wells at the river is perceived as clean after they pour out the brown, dirty water. The borehole water is hard. If the water appears brown, it may be boiled or treated with chlorine, but this is rare. Participants perceived dam water to be dirty because elephants, goats, and livestock drink from the source and leave waste in it. Dam water may be treated using a sieve or, if they have access, chlorine. Participants reported treating water from other sources with chlorine or boiling only if the water appears to be brown. | Water quality from sources other than dams was generally perceived to be good. Women reported that their water is clear with no taste in the rainy season, unless it is from the small dams. Water from dams was noted to be brown. At the dug wells at the river, they poured out the dirty water and only collected the clean water, but one participant reported that sometimes she just gets dirty water and goes home. Participants noted that water may taste salty in the dry season. One river in particular was identified as the cleanest, but it was the farthest away from the community. One participant reported treating water every time unless it is from the borehole. Other participants treat only the water from the dam with chlorine, if they have been provided with chlorine recently. If chlorine packets are expired, they will use the water for washing clothes and bathing but not for drinking. | Women reported that water from the borehole is clean. When they collect water from dug wells at the river, they scooped out the brown water and only collected the clear water. They did report contamination of sources from wild animals, and that in the dry season, sometimes the water is brown and has a taste and "bad vapours." One participant reports always treating water from the river, but not from the borehole. Water treatment is dependent on having access to chlorine. |

| **S1d Table. Zimbabwe** | | | | | | |
| --- | --- | --- | --- | --- | --- | --- |
| **Community** | **1 n=4** | **2 n=4** | **3 n=6** | **4 n=2** | **5 n=3** | **6 n=14** |
| **Water Sources Visited During Data Collection** | One participant journeyed to an unprotected dug well at a river/stream; the other three journeyed to a dam where they collected surface water. | All participants collected water from an unprotected dug well at the river. | Participants collected water from a borehole, a protected well at a dam, and an unprotected dug well at a stream. | Participants collected water from a tap and a borehole. | Participants collected water from an unprotected dug well in a river/stream or from a borehole. | Participants collected water from a borehole, a tap, or an unprotected dug well at a river/stream. |
| **Other Water Sources Used by Participants, But Not During Data Collection** | Other water sources mentioned included rivers or streams, surface water at a dam, or rainwater harvesting from rooftops and water "paddles" during the rainy season. One participant reported collecting from JoJo tanks for a period of 2 weeks, but was then told to wait for an official opening to use this source again. | Other water sources mentioned included three different rivers, depending on the season, small streams near the house during the rainy season, a rock with a dug basin, and a "trophy" which pipes water from the river that is accessible by handpump. | Other water sources mentioned included surface water at the dam, a protected well covered in a slab behind the dam, pondwater, and rainwater harvesting from rooftops and streams during rainy season. One participant reported occasionally paying someone to collect water for her. | Other water sources mentioned included at tap installed at the local school, an open well dug as a community, a borehole, and a stream. | Other water sources mentioned included an unprotected well and rainwater harvesting from the roof or surface water during the rainy season. | Other water sources mentioned included a borehole, a tap installed at school, surface water collection or rainwater harvesting from rooftop in rainy season, several different unprotected wells at rivers or streams, unprotected wells at homesteads, and water "paddles" from dwalas (large rocks with crevices that accumulate water). Participants that used the borehole or tap as a source for drinking water reported that the water was rationed, so they also visited additional sources for uses other than drinking water, such as bathing and washing clothes. |
| **Method of collection** | The participant that collected water from the dug well used sand abstraction, digging next to or in a riverbed until she hit water. She then drew the first layer of water out and then waited for the water level to rise. Participants at the dam scooped water out directly. | Participants used sand abstraction. They dug next to or in the riverbed until they hit water. They scooped out the first layer of water and then waited for clean water to rise, using a scooping plate or cup to get water from the well into the buckets. When finished, they covered the pit to protect it from donkeys. Sand abstraction. Marks spot with bucket. Draws water by removing first layer from unprotected well at stream and then waiting for clean water level to rise. | Women pumped water at the borehole with help from others, bent and drew water from a well using tins or plastic containers, stepped into water at a dam to fill a bucket, or collected water via sand abstraction by digging into a hole in or near a riverbed, and then scooping out water. | (Not described) | Women collected water by pumping at the borehole with the help of two other women, or via sand abstraction, digging in or next to a river until water level rises and scooping water out. | Women collected water via sand abstraction. They dug holes in or near rivers, scooped water from the surface using a plate, and then placed a drum inside the abstraction which then filled with water. The depth of the hole sometimes required someone to kneel on the ground with their head hanging inside the drum to collect water. Water was also collected by pumping at a borehole with the help of others, or by drawing water from a well using a rope. |
| **Number and timing of trips** | Most women reported two trips per day, one in the morning and one in the evening, with a possible third or fourth trip. Participants noted that the schedule may be adjusted during the farming season or vary between rainy and dry season. To avoid queues, they may collect early in the morning. One participant reported that she collects twice a day if she is using a wheelbarrow to transport water but otherwise collects 3 times a day. | Most participants reported going 2 or 3 times a day. One reported taking 5 trips a day in the dry season. All participants reported collecting water in the morning and evening; two noted that if they collected twice in the morning, they would not always go in the evening. One participant stated that in addition to water collection, she went to the river twice a week to wash clothing. Another noted that sometimes she is unable to collect water because of other commitments such as farming or attending funerals or meetings, so she collects extra water the day before. | Women reported a variety of different times for when they collected water. One woman reported going to the dam as many as 6 times in a day and going to the borehole 3-6 times a day. Another reported collecting from the borehole twice a day, in the morning and later in the afternoon, but being limited by set times enforced on when the borehole can be used. Another reported one trip a week to the stream. Two participants changed how often they collected based on if they transported water with assistance from a wheelbarrow or ox-drawn cart, or if they were accompanied by someone. One woman noted that morning collection times may be affected by funerals or community programs, such as trainings or development projects. When water supply is low, participants report waking up as early as 4 A.M. because they may have to queue for several hours. | One participant reported going twice a day to the tap and three times a day to the well, beginning at 5:30 in the morning. The other reported going three times a day to the borehole and twice a day to the stream, beginning at 7 in the morning, and noted that people are already queuing when she arrives | Two participants reported collecting twice a day in the dry season, in the morning and late afternoon, but one of the two said she only goes to the source once a day during the rainy season. Another participant reported collecting four times -- twice to the borehole and twice to the stream, also in the morning and late afternoon. | Most participants reported going to a water source 2 or 3 times a day and mentioned that there are frequently waits or queues at the water sources. The number of times and the timing of the visits that they reported varied based on if a participant has visitors, meaning her household needs more water, if she has visitors that she can't leave alone to go collect water, if children or other family members assist with carrying water, if they have additional sources during the rainy season, if there are other responsibilities such as farming to attend to, or if the tap or borehole is closed when they arrive. One participant reported going three times in the morning but only twice on Sundays. Another reported going to the borehole three times a week and the river three times a day, with long queues and waits at the river. Another reported going to the borehole three times a day but does not go to the river every day. |
| **Method of carriage** | Women carried water using a wheelbarrow or by carrying buckets on their heads. | Women carried water in buckets on their heads. Women at the water point assisted each other in placing buckets on heads. Two participants reported occasionally using scotch carts, if neighbors could share or there were donkeys available. Others at the water point may assist in putting buckets on heads. | Participants carried 20 and 25 liter buckets on head. Two participant reported often using wheelbarrows, which can be used to carry more than one bucket, but it was not documented whether they were using them on this water journey. Another reported occasionally being assisted by her husband or aunt and a donkey-drawn cart. | Women carried containers on their heads. | All participants carried water in 20 L buckets. Two methods of carriage were not specified; one participant carried water on her head. | Participants carried water in buckets, usually at least one bucket of 20 liters. Sometimes additional, smaller buckets are carried, either by the participant or by a child. Buckets were carried on the head or in sctoch carts. If they used a scotch cart, they were able to carry additional buckets. One participant reported having a scotch cart, but she only uses it with her neighbors or son as she is unable to use it alone. Another participant reported that her husband sometimes helps her to use a scotch cart. |
| **Terrain and risks** | The terrain was bumpy, with hills, loose stones. Participants reported that it is hard to climb the hills when carrying buckets of water or pushing a wheelbarrow. One participant reported that she has to change her route in the rainy season because her path becomes farmland and is closed. Participants reported that there are snakes. Participants reported that children can drown or be hurt playing at the dam. The road to the river was better, as there is a small path, but there were sandy stretches that are hard to walk or push wheelbarrows on. | The terrain was rocky and steep, with thorny trees, roots, and loose stones, where women trip and fall. One participant reported having fallen and broken a full water bucket. There are snakes. In the dry season, there are large pits where participants report a person can be buried by sand. Participants also noted animals including snakes and jackals and hyenas that eat domestic animals. Participants reported having heard of elephants passing but not encountering them. Women reported that they collect water with neighbors because it is not safe to go alone. They feared attacks from people. They also noted that at one point, there was a rapist with a machete who attacked two women. He was caught, but they were still frightened.   p1 - afraid that children will fall in and drown | The pathway was narrow and uneven, with steep ascents and sand stretches, as well as thorns, bushes, and loose stones; it was easy to trip and difficult to push a wheelbarrow. There were dwalas, or large rocks, that women had to climb up and down, and they reported being afraid of falling. Participants also reported being afraid of dangerous people. They noted that sometimes there are traditional healers performing rituals at the dam, and there is a snake in the river and on the path to the borehole. | One pathway had rocky, mountainous stretches, steep slopes, and bushy areas. There were sandy stretches which participants reported were challenging to walk on while carrying water, and the sand becomes hot in the sun. The main road had a lot of motorbikes, and participants reported concerns about children being hit. One participant noted that the pathway they were using was in a field, so in the rainy season they have to find another route, so they do not cut through someone's crops. | The terrain was mountainous, with rocky, thorny paths, tripping hazards, sandy stretches which may bury shoes, riders on bikes speeding on the way, and steep slopes. There were loose stones. Participants reported that baboons may attack goats and cows. | The terrain was rough, with steep slopes, dried streams, loose stones, bushy areas, and thorns. On the way to the borehole, there were narrow stretches where two people could not walk side by side. Participants reported that sandy stretches in the road become hot and it's hard to walk on them, especially while carrying a load of water and a baby. Participants noted that there are hyenas as well as bees at the borehole and a snake in the river. A grandmother once fell into the stream and had to be rescued. Participants noted reports of scolding, abuse, and harassment of women and girls. One participant noted that they pass through other people's farms to get to the water source |
| **Physical effects of water collection** | Women reported experiencing whole body numbness and tired arms. One participant reported that the tiredness was worse when she used a wheelbarrow because the weight of water is greater and it's difficult to push the wheelbarrow. | Women reported tiredness and pain in the neck, back and knees. One participant noted the difficulties of water collection because of her diabetes and asthma. | Women reported chest pains and shortness of breath. One participant noted that the journey is not too far, but that the borehole requires a lot of energy to pump. | A participant reported chest pains from water collection. | Women reported back pain and overstraining the body. | Women reported numbness of the legs and body aches and pains in the back, and knees from carrying water. Chest and shoulder pain were noted as a result of pumping at the borehole. |
| **Children** | Participants reported that if young children are not in school, they may accompany women in water collection, carrying smaller or partially full buckets. Older children may fetch on their own if they don't have school. One woman carried an infant on her back and noted that she sometimes lets the child sit in the wheelbarrow on the journey to the water source; she also often collects with her three adolescent sons, who help her carry water back. | One participant carried her child and has to breastfeed on the journey. Two participants report that children may go collect water on weekends. Another reported that children are not allowed to collect water. | Participants reported varying information regarding the presence of children on the water journey. One participant reported that her children may go to fetch water if she isn't feeling well. Most participants reported that children do not go alone to most water sources because of safety concerns. One participant stated that children in the community are not allowed to go if they are under 18. One participant reported that her children can go alone to the pond, which is near the home, but she is hesitant to send her boys to fetch water at the dam because they did not pay attention to water quality. Only one participant was observed to have her child, an older son, with her. | One participant reported sometimes going with her 6-year old, who carries a partially full 10-liter container. It was not documented if participants had children with them on this journey. | All participants carried babies with them on their water journey. One participant dropped a child off part of the way to be minded at a shop. | Two participants were observed carrying babies on this journey, and more participants reported that they often carry babies or bring their children. Participants also reported going with older children before school, bringing children along and having to walk slowly to accommodate them, leaving small children at home with older children, leaving children alone, or having challenges in being unable to find childcare, which may cause delays in going to borehole. |
| **Other work done during water collection** | Participants reported or were observed washing clothes at the water source, picking fruit when in season, and chopping firewood. The participant who reported chopping firewood leaves it along the way and then goes back to collect it after she has dropped the water at her house. | Participants reported or were observed watering gardens, washing clothes in the river, collecting firewood on the way and going back for them, harvesting mopane worms and gathering fruit in the proper season, and rinsing out buckets and lids for water collection. One participant reported that bathing is not permitted in the river so she washes at the dam or at home; another reported that bathing is permitted, but only if the water is collected and carried to nearby trees to avoid contaminating the water source. | Participants reported or were observed gathering fruit when in season, fetching firewood, washing buckets and clothes, and driving cattle and goats to source to drink. One participant, when she collects firewood, leaves it on the way and then returns to collect it. | One participant reported gathering firewood and doing laundry while at or going to the water source. | Participants reported or were observed cleaning buckets, washing clothes, fetching firewood to collect later, driving cattle to the stream for drinking, and picking fruits when in season. One participant watered vegetable gardens on the other side of the river, making up to 8 trips back and forth, and carried vegetables back. | Participants reported or were observed cleaning buckets, washing clothes, gathering fruit when in season, driving cows to the river, watering and carrying manure to a garden, and carrying vegetables back from the garden. One participant reported visiting a shop she has on the way to check on her sales. |
| **Perceived water quality and water treatment** | Participants reported that the dam water is unsafe/contaminated, as animals drink from the water. Observers confirmed that the dam appeared heavily polluted. One participant reported that well water is safer than dam water. No one reported treating water. | Women reported several concerns over water quality. One said that in the rainy season, the stream is not safe because community members soil and urinate in the water and may throw diapers in the area. Another participant reported water from the creeks is clean and that is what they drink, and that the river water is usually clean, but sometimes there are red insects in the water, so she boils the water and transfers it to another bucket. One participant reported that the metal drum inserted for water collection is rusty, which contaminates the water and leads to diseases. She also noted that sometimes the drum is found without the cover and animals are drinking from the stream, so she uses a well at another stream nearby without a drum, and covers it with sand to keep it unpolluted, but the water still is not clean. | Perceptions of water quality varied by source. Participants reported that dam water is dirty, as animals drink from the dam. Participants did not treat water from the borehole, and some participants also said that roof water is safe to drink, whereas one participant thought sand filtration resulted in cleaner water. One participant reported that water from the well is safer than water from the dam, but others said that water from the well is not fully protected or safe for drinking. Reported methods for treating water included using pouring water through a cloth to absorb dirty particles, boiling water, and using water treatment tablets. | One woman reported that water is safe and there is no need to boil it | One participant reported water quality concerns, saying that the water is rusty from the borehole, and there are worms in the stream. One participant reported keeping buckets off the floor so the collected water stays cleaner. No participants reported treating water, though one participant noted that chlorine tablets could be acquired at the health mission. | Many of the participants used to treat water using purification tablets, but one reported that her family didn't like the taste and others reported that they no longer have water tablets. Two participants reported that boiled water is tasteless or does not taste good, and one said that she is too tired to boil it most of the time. Some participants reported that water from some sources is clean and does not need treatment, particularly from the stream, the tap, and the borehole. Many participants reported treating water from sources other than the borehole, which they perceive to be clean, though one participant said that the borehole water looks rusty. Two participants report that if they are collecting water from the river, they will treat by boiling it or using water tablets. Participants did not report treating any water not used for drinking. Reported steps taken to keep water clean tend to be storing the water in closed buckets with a lid, setting the drinking water aside from the rest of the water for safekeeping, and being careful to only use one cup to fetch water from the bucket of drinking water to avoid contamination. |
