## Supplementary material for "Women’s experiences collecting and accessing water in Guatemala, Honduras, Kenya, and Zimbabwe: A mixed-methods investigation": S1 Tools

**A mixed-methods investigation of water burden: Supplement 1 Tools**

Bethany A. Caruso^1*^, Thea Mink^1^, Madeleine Patrick^1^, Emily Ogutu^2^, Cameron Dawkins^1^, Olivia Bendit^1^, Mahnoor Fatima^1^, Ingrid Lustig^1^, Alicia Macler^1^, Jera White^1^, Alondra Zamora^1^, Alberto Emanuel Santos López^3^, Héctor Salvador Peña Ramírez^3^, Carlos Daniel Sic^3^, Jorge Lemus Chávez^4^, Sandra Antonio^5^, Jazmina Nohemí Irías^5^, Gladys Ramos^6^, Everlyne Atandi^7^, Peter Mwangi^7^, Peter Koome^8^, Rohin Otieno Onyango^8^, Petronilla Andiba Otuya^8^, Paul Ruto^8^, Morris Chidavaenzi^9^, Jammaine Jimu^9^, Sithandekile Maphosa^9^, Makaita Maworera^9^, Munyaradzi Damson^10^, Sheela S. Sinharoy^1^

^1^ Hubert Department of Global Health, Rollins School of Public Health, Emory University, Atlanta, GA, USA

^2^ Gangarosa Department of Environmental Health, Rollins School of Public Health, Emory University, Atlanta, GA, USA

^3^ World Vision Guatemala, Guatemala City, Guatemala

^4^ Centro Universitario de Occidente, de la Universidad de San Carlos de Guatemala, Quetzaltenango, Guatemala

^5^ World Vision Honduras, Tegucigalpa, Honduras

^6^ Universidad Nacional Autónoma de Honduras, Tegucigalpa, Honduras

^7^ World Vision Kenya, Nairobi, Kenya

^8^ St. Paul’s University, Nairobi, Kenya

^9^ World Vision Zimbabwe, Harare, Zimbabwe

^10^ Datalyst Africa, Harare, Zimbabwe

* Corresponding author

 (BAC)

**S1 Tools.** Data collection tools for participant demographic information, go-along interviews, and semi-structured observations

**S1a Tools.** Participant Demographic Information

**S1b Tools.** Water Journey Go-along Interview Guide

**S1c Tools.** Water Journey Semi-structured Observation Guide

**S1a Tools.** Participant Demographic Information

| **A. Activity information** | | | |
| --- | --- | --- | --- |
| **A001** | Participant ID#: __________________________________ | **A005** | Date: (y/d/m) __ __ __ __ / *__ _**_ / __ __* |
| **A010** | Community Name: __________________________________ | **A015** | Community ID#: ____________________________________ |
| **A017** | **Select activity person will participate in:** | ☐ 1. Key Informant Interview  ☐ 2. Water Journey Go-Along Interview  ☐ 3. FGD with Women  ☐ 4. FGD with Men | |
| **A18** | **If there is more than one activity of this type today, indicate activity number:** | ☐ 1. Not applicable; only activity of this kind here today  ☐ 2. Activity number: **__ __** | |
| **A020** | **Activity Start time:** __ __ : __ __ pm / am | **A025** | **Activity End time:** __ __ : __ __ pm / am |
| **A030** | **Person Filling** **form:__________________** | **A040** | **Consent Obtained:** ☐ 1. Yes ☐ 2. No |

| **B. Participant Demographic Information** | | | |
| --- | --- | --- | --- |
| **D01** | **Participant Gender**  ☐ 1. Woman ☐ 2. Man | **D02** | **Participant Age __ __**  *(If younger than 18, participant ineligible. End.)* |
| **D03a** | **What is your marital status?**  ☐ 1. Single, never been married  ☐ 2. Unmarried, but have partner  ☐ 3. Married  ☐ 4. Separated  ☐ 5. Divorced  ☐ 6. Widowed | **D03b** | **What type of family structure do you have? [Kenya]**  ☐ 1. Monogamy  ☐ 2. Polygamy |
| **D04** | **What is the highest level of school you completed?**  ☐ 1. Never attended school  ☐ 2. Completed some Primary  ☐ 3. Completed Primary  ☐ 4. Completed Secondary  ☐ 5. Completed schooling above Secondary | | |
| **D05** | **How many people (including yourself) live in your household? __ __**  [Usually number of people sharing meals] | **D06** | **How many children live in your household?**  **__ __**  [This is the total number under age 18.] |
| **D07a** | **Some people take up jobs for which**  **they are paid in cash or kind. Others sell things, have a small business or work on the family farm or in the family business. In the last** **30 days, have you done any of these things or any other work?** ☐ 1. Yes ☐ 2. No | **D07b** | ***If participant has engaged in work in the past 30 days (D07a=yes):***  **What are the activities you have done for work?** (write answer) |
| **D08a** | ***If married,* In the last** **30 days, has your spouse [husband/wife] done any of these things or any other work?**  ☐ 1. Yes ☐ 2. No | **D08b** | ***If participant’s spouse has engaged in work in the past 30 days (D08a=yes):***  **What are the activities your spouse has done for work?** (write answer) |
| **D09a**  **D09b** | **What is the primary source of DRINKING water used by members of the household in the past 7 days / week?**  **What is the primary source of water FOR OTHER NEEDS used by members of the household in the past 7 days / week?**  **(includes for cooking, bathing, washing clothing, etc.)** | **D09a D09b**  **Drinking Other Uses**  ☐ ☐ 01. No other source used  ☐ ☐ 10. Piped Water  ☐ ☐ 21. Tube well/Borehole  ☐ ☐ 31. Protected Dug Well  ☐ ☐ 32. Unprotected Dug Well  ☐ ☐ 41. Protected Spring  ☐ ☐ 42. Unprotected Spring  ☐ ☐ 51. Rainwater  ☐ ☐ 61. Tanker Truck  ☐ ☐ 71. Cart with small tank  ☐ ☐ 72. Water Kiosk  ☐ ☐ 81. Surface Water *  ☐ ☐ 91. Packaged bottled water  ☐ ☐ 92. Packaged Sachet water  ☐ ☐ 96. Other_____________ *River, dam, lake, pond, stream, canal, irrigation channel | |
| **D10a** | **Where is the household’s main drinking water source?**  ☐ 1. Source in dwelling  ☐ 2. Source in yard/plot  ☐ 3. Source elsewhere (beyond yard/plot) | **D10b** | **How long does it take for members of your household to go to the drinking water source, get water, and come back?**  ☐ 1. __ __ __ minutes  ☐ 88. Do not know  ☐ 99. Not applicable; do not collect drinking water /source on property |
| **D10c** | **How many days in a week does your household collect water from the primary drinking water source?**  __ __  Enter 1-7 for number of days per week  Enter 99 if not applicable  Enter 88 if do not know | **D10d** | **Who is *primarily responsible* for collecting drinking water for the household?**  ☐ 1. Respondent  ☐ 2. Adult woman (age 18 or over)  ☐ 3. Girl (under age 18)  ☐ 4. Adult man (age 18 or over)  ☐ 5. Boy (under age 18)  ☐ 99. Not applicable; do not collect drinking water |
| **D11** | **In the last month, has there been any time when your household did not have** **sufficient quantities of drinking water when needed?**  ☐ 1. Yes, at least once  ☐ 2. No, Always sufficient  ☐ 88. Do not know | **D11a** | ***If the source for other uses is different than the drinking water source,***  **Where is the household’s main water source for *other uses*?**  ☐ 1. Source in dwelling  ☐ 2. Source in yard/plot  ☐ 3. Source elsewhere (beyond yard/plot) |
| **D11b** | ***If the source for other uses is different than the drinking water source,***  **How long does it take for members of your household to go to the water source for other uses, get water, and come back?**  ☐ 1. __ __ __ minutes  ☐ 88. Do not know  ☐ 99. Not applicable; do not collect drinking water /source on property | **D12** | **Who is *primarily responsible* for collecting water for other uses for the household?**  ☐ 1. Respondent  ☐ 2. Adult woman (age 18 or over)  ☐ 3. Girl (under age 18)  ☐ 4. Adult man (age 18 or over)  ☐ 5. Boy (under age 18)  ☐ 99. Not applicable; do not collect water for other uses |
| **D13a** | **Does your household have access to a toilet facility?**  ☐ 1. Yes, in dwelling  ☐ 2. Yes, in yard/plot  ☐ 3. Yes, elsewhere (beyond yard/plot)  ☐ 4. No household access to a toilet facility | **D13b** | **If your household has access to a toilet facility, do you share it**?  ☐ 1. No, not shared with other households  ☐ 2. Yes, shared with specific households  ☐ 3. Yes, shared with public/community  ☐ 99. Not applicable; no access to a toilet facility |

| **C. Water Journey Information**  *Only collect from Water Journey Participants.* | | | |
| --- | --- | --- | --- |
| **WD01** | **Height:** _ _ _ cm | **WD02** | **Weight: _ _** kg |
| **WD03** | **Date of birth** (yyyy/dd/mm): _ _ _ _ / _ _ / _ _ | | |
| **WD10** | **Length of time lived in community:**  **_ _** years | **WD11** | **Do you intend to leave the community in the next year?**  ☐ 1. Yes  ☐ 2. No  ☐ 88. Do not know |

**S1b Tools.** Water Journey Go-along Interview Guide

A. **Background information**

| **Guide: Go-along Interviews and Observations** | | | |
| --- | --- | --- | --- |
| **A010.** | Community Name: __________________________________ | **A015.** | Community ID#: ___________________________________ |
| **A020.** | Activity Start time: __ __ : __ __ pm / am | **A025.** | Activity End time: __ __ : __ __ pm / am |
| **A055.** | Date: (y/d/m) __ __ __ __ / *__ _**_ / __ _* | **A040.** | Consent Obtained: ☐ 1. Yes ☐ 2. No |
| **A045.** | Recorder ID:_______________________ | **A046.** | Recording #__________________________ |
| **A030.** | Interviewer: __________________________________ | **A035.** | Note Taker/Observer: ____________________________________ |
| **Introduction** | | | |

B. **Quantitative metrics**

Take the weights of any kind of load that women carry while transporting water. This should include weights of laundry when women do their laundry at the water source.

| **Items** | **Weight (kg)** |
| --- | --- |
| Water containers without water |  |
| Water container with water |  |
| Child(ren) (if carried by participant) |  |
| Laundry (dry) |  |
| Laundry (wet) |  |
| Other (specify): |  |
| Other (specify): |  |
| Other (specify): |  |

C. **Interview questions**

| **Pre and post water collection**   1. Can you walk me through your typical day in relation to water collection? | **Probe:**  What preparations do you make before going to collect water?  **How long does it typically take you to collect water?** |
| --- | --- |
| 2. Where are you currently walking to collect water? | **Probe:**  -What do you plan to use this water for? |
| 3. How often do you collect water for household use at this location (times/week)? |  |
| 4. Where else do you collect water from? | **Probe:**  -Drinking water  -Water for other uses  **Probe**: Laundry, bathing, other  -If water is collected from multiple sources: How do you decide where to collect water from for different activities?  **Probe**: Distance, time, weight, water quality |
| 5. Do you collect water for animal use as well? |  |
| 6. How many trips do you make in a day for water collection? | **Probe:**  -Does this change with seasonality?  -Does this change from day-to-day based on household use (e.g. laundry and other chores that may not be done on a daily basis). |
| 7. What time(s) of the day do you normally go to collect water? | **Probe:**  -Why that time?  -Is there a specific time of the day that you cannot go to the water source? Why?  -If water is collected from multiple sources: repeat question based on source |
| 8. Do you go to the water source by yourself or in a group? Why? | **Probe:**  Do children also go to collect water from these sources? If not, why? |
| 9. Other than water collection, what other activities are you involved in while going to collect water, either along the way or at the water source? | **Probe** (Do not ask probes as yes/no questions; allow participants to describe experiences):  -Socializing with other women  -Fetching firewood on the way  -Picking fruits or vegetables  -Bathing (themselves / children) at the water source  -Washing dishes / laundry at the water source |
| 10. What activities are you involved in to clean or store the water once it has been collected? | **Probe** (Do not ask probes as yes/no questions; allow participants to describe experiences):  -Boiling water  -Filtering water  -Storing water for future use |
| 11. If a water source was brought nearby, how would that change how you manage your time? | **Probe:**  -Productive activities (e.g., garden / agriculture)  -Income-generating activities (e.g., start a business)  **-A**bility to rest |
| 12. What are some of the challenges you experience while collecting water? | **Probe** (Do not ask probes as yes/no questions; allow participants to describe experiences):  -Water scarcity  -Attacks by wild animals  -Attacks by people  -Falls due to terrain  -Having to left heavy containers  -Pain  -Finding childcare or watching children while collecting  -Long lines  -Difficulty walking long distances |
| 13. Do you leave children or other dependents at home while you collect water? | **Probe**   - Number of children/dependents - Frequency - Do they have supervision? |

**S1c Tools.** Water Journey Semi-structured Observation Guide

**Time tracking**

For each activity, document what time the activity begins and ends. Include when the water journey begins, any rest taken along the way, when they arrive at a water point, how long the labor of collection takes, if there is any rest at the water point, when they leave the water point, and when they arrive back at their house.

Some activities, like taking a break, may occur multiple times on the water journey. Document each time it happens. If there are other activities not listed here, make sure you make a note of those as well.

Example:

| **Start Time** | **End Time** | **Activity** |
| --- | --- | --- |
| 8:05 AM | 9:50 AM | Leaves and walks to water collection point |
| 10:50 AM | 11:05 AM | Rests and talks to other women at water point |
| 11:05 AM | 11:45 AM | Pumps water |
| 11:45 AM | 12:00 PM | Rests, feeds child |
| 12:00 PM | 1:30 PM | Walks home with water |
| 1:35 PM | 1:40 PM | Rests, takes drink |
| 1:40 PM | 2:20 PM | Walks home with water |

| **Start Time** | **End Time** | **Activity** |
| --- | --- | --- |

**Qualitative observation**

**Note to observer:** Please document your observations of the water collection journey. We have suggested specific things to pay attention to below, but they are not exhaustive nor are they in order, so please do not limit your observations to what is below. Take notes on the entire journey as you go on it; these notes can be in shorthand as after the observation, you will write out additional notes you did not have time for and debrief with the team. Feel free to also draw maps and pictures as relevant.

| **Observation** | **Detailed description** |
| --- | --- |
| Type and description of containers women use for carrying water | Describe containers (i.e., jerry cans, buckets)  Number of containers  Volume  How the water containers are carried (on head, by handle)  Any other means of transporting water e.g., use of animals like donkeys |
| Health condition and additional items that the woman carry/ bring along to the water source | For example:  Pregnancy  Any visible disability  Carrying a child (i.e., on her back)  Travelling with small animals |
| Terrain | For example:  Hilly, flat, muddy, dry, etc. |
| Water collection process | Describe water access point(s)  Describe water access behavior  When does water collection start, and when does it end?  Who else is at the water source?  Are there are specific areas within the water source specifically meant for women (only women collect water from that point)? |
| Any other activity that the woman is engaged in while collecting water | For example:  Taking a break/ rest  Eating  Drinking  Feeding child(ren)  How long are activities? |
| Other observations. Any encounter with wild animals, injuries, encounters with other people |  |
