## Supplementary material for "Women’s experiences collecting and accessing water in Guatemala, Honduras, Kenya, and Zimbabwe: A mixed-methods investigation": S2 Table

**A mixed-methods investigation of water burden: Supplement Table 2**

Bethany A. Caruso^1*^, Thea Mink^1^, Madeleine Patrick^1^, Emily Ogutu^2^, Cameron Dawkins^1^, Olivia Bendit^1^, Mahnoor Fatima^1^, Ingrid Lustig^1^, Alicia Macler^1^, Jera White^1^, Alondra Zamora^1^, Alberto Emanuel Santos López^3^, Héctor Salvador Peña Ramírez^3^, Carlos Daniel Sic^3^, Jorge Lemus Chávez^4^, Sandra Antonio^5^, Jazmina Nohemí Irías^5^, Gladys Ramos^6^, Everlyne Atandi^7^, Peter Mwangi^7^, Peter Koome^8^, Rohin Otieno Onyango^8^, Petronilla Andiba Otuya^8^, Paul Ruto^8^, Morris Chidavaenzi^9^, Jammaine Jimu^9^, Sithandekile Maphosa^9^, Makaita Maworera^9^, Munyaradzi Damson^10^, Sheela S. Sinharoy^1^

^1^ Hubert Department of Global Health, Rollins School of Public Health, Emory University, Atlanta, GA, USA

^2^ Gangarosa Department of Environmental Health, Rollins School of Public Health, Emory University, Atlanta, GA, USA

^3^ World Vision Guatemala, Guatemala City, Guatemala

^4^ Centro Universitario de Occidente, de la Universidad de San Carlos de Guatemala, Quetzaltenango, Guatemala

^5^ World Vision Honduras, Tegucigalpa, Honduras

^6^ Universidad Nacional Autónoma de Honduras, Tegucigalpa, Honduras

^7^ World Vision Kenya, Nairobi, Kenya

^8^ St. Paul’s University, Nairobi, Kenya

^9^ World Vision Zimbabwe, Harare, Zimbabwe

^10^ Datalyst Africa, Harare, Zimbabwe

* Corresponding author

 (BAC)

| **S2 Table (part a).** Water Journey estimated time comparison and discordance, by country (n=36) | | | | | | | | | | | | | |
| --- | --- | --- | --- | --- | --- | --- | --- | --- | --- | --- | --- | --- | --- |
|  | **Guatemala** | | | | | |  | **Honduras** | | | | | |
|  | Any | | Drinking | | Other uses | |  | Any | | Drinking | | Other uses | |
|  | n | % | n | % | n | % |  | n | % | n | % | n | % |
| **Total Comparisons^1^** | 4 | | 3 | | 1 |  |  | 5 |  | 3 |  | 2 |  |
| **Time** |  |  |  |  |  |  |  |  |  |  |  |  |  |
| ***Underestimates*** | **1** | **25.0** | **0** | **0.0** | **1** | **100.0** | | **0** | **0.0** | **0** | **0.0** | **0** | **0.0** |
| ≥60 minute underestimate | 1 | 25.0 | 0 | 0.0 | 1 | 100.0 | | 0 | 0.0 | 0 | 0.0 | 0 | 0.0 |
| 30-59 minute underestimate | 0 | 0.0 | 0 | 0.0 | 0 | 0.0 | | 0 | 0.0 | 0 | 0.0 | 0 | 0.0 |
| 16-29 minute underestimate | 0 | 0.0 | 0 | 0.0 | 0 | 0.0 | | 0 | 0.0 | 0 | 0.0 | 0 | 0.0 |
| ***Estimate +/- 15 minutes of actual time*** | **3** | **75.0** | **3** | **100.0** | **0** | **0.0** | | **5** | **100.0** | **3** | **100.0** | **2** | **100.0** |
| ≤15 minute underestimate | 3 | 75.0 | 3 | 100.0 | 0 | 0.0 | | 3 | 60.0 | 3 | 100.0 | 0 | 0.0 |
| ≤15 minute overestimate | 0 | 0.0 | 0 | 0.0 | 0 | 0.0 | | 2 | 40.0 | 0 | 0.0 | 2 | 100.0 |
| ***Overestimates*** | **0** | **0** | **0** | **0.0** | **0** | **0.0** | | **0** | **0.0** | **0** | **0.0** | **0** | **0.0** |
| ≥60 minute overestimate | 0 | 0.0 | 0 | 0.0 | 0 | 0.0 | | 0 | 0.0 | 0 | 0.0 | 0 | 0.0 |
| 30-59 minute overestimate | 0 | 0.0 | 0 | 0.0 | 0 | 0.0 | | 0 | 0.0 | 0 | 0.0 | 0 | 0.0 |
| 16-29 minute overestimate | 0 | 0.0 | 0 | 0.0 | 0 | 0.0 | | 0 | 0.0 | 0 | 0.0 | 0 | 0.0 |
| **Discordance** |  | |  | |  | |  | |  | |  | |  |
| ***No time discordance*** | **1** | **25.0** | **1** | **33.3** | **0** | **0.0** | | **1** | **20.0** | **1** | **33.3** | **0** | **0.0** |
| Estimated and actual both ≤ 30min | 1 | 25.0 | 1 | 33.3 | 0 | 0.0 | | 1 | 20.0 | 1 | 33.3 | 0 | 0.0 |
| Estimated and actual both > 30min | 0 | 0.0 | 0 | 0.0 | 0 | 0.0 | | 0 | 0.0 | 0 | 0.0 | 0 | 0.0 |
| ***Time discordance*** | **3** | **75.0** | **2** | **66.7** | **1** | **100.0** | | **4** | **80.0** | **2** | **66.7** | **2** | **100.0** |
| Estimate at or under 30min, actual over 30min | 2 | 50.0 | 2 | 66.7 | 0 | 0.0 | | 2 | 40.0 | 2 | 66.7 | 0 | 0.0 |
| Estimate over 30min, actual at or under 30min | 1 | 25.0 | 0 | 0.0 | 1 | 100.0 | | 2 | 40.0 | 0 | 0.0 | 2 | 100.0 |
| ^1^ Data from 10 women were excluded because they did not collect water to bring home (9 Guatemala, 1 Kenya) and from 10 women because they did not provide estimated times (5 Guatemala, 1 Honduras, 4 Zimbabwe). Data from 38 women were excluded because their water source type was not the same for their estimated and measured water journey times (5 Guatemala, 10 Honduras, 9 Kenya, 14 Zimbabwe). For the Guatemala participant who completed two water journeys, only her water journey to collect water was included. ￼ | | | | | | | | | | | | | |

| **Table S2 (part b).** Water journey estimated time comparison and discordance, by country (n=36) | | | | | | | | | | | | | | | | | | | | |
| --- | --- | --- | --- | --- | --- | --- | --- | --- | --- | --- | --- | --- | --- | --- | --- | --- | --- | --- | --- | --- |
|  | **Kenya** | | | | | |  | **Zimbabwe** | | | | | |  | **Total** | | | | | |
|  | Any | | Drinking | | Other uses | |  | Any | | Drinking | | Other uses | |  | Any | | Drinking | | Other uses | |
|  | n | % | n | % | n | % |  | n | % | n | % | n | % |  | n | % | n | % | n | % |
| **Total Comparisons^1^** | 12 |  | 12 |  | 0 |  |  | 15 | | 12 |  | 3 |  |  | 36 |  | 30 |  | 6 |  |
| **Time** |  |  |  |  |  |  |  |  |  |  |  |  |  |  |  |  |  |  |  |  |
| ***Underestimates*** | **2** | **16.7** | **2** | **16.7** | **0** | **0.0** | | **1** | **6.7** | **1** | **8.3** | **0** | **0.0** | | **4** | **11.1** | **3** | **10.0** | **1** | **16.7** |
| ≥60 minute underestimate | 2 | 16.7 | 2 | 16.7 | 0 | 0.0 | | 0 | 0.0 | 0 | 0.0 | 0 | 0.0 | | 3 | 8.3 | 2 | 6.7 | 1 | 16.7 |
| 30-59 minute underestimate | 0 | 0.0 | 0 | 0.0 | 0 | 0.0 | | 0 | 0.0 | 0 | 0.0 | 0 | 0.0 | | 0 | 0.0 | 0 | 0.0 | 0 | 0.0 |
| 16-29 minute underestimate | 0 | 0.0 | 0 | 0.0 | 0 | 0.0 | | 1 | 6.7 | 1 | 8.3 | 0 | 0.0 | | 1 | 2.8 | 1 | 3.3 | 0 | 0.0 |
| ***Estimate +/- 15 minutes of actual time*** | **1** | **8.3** | **1** | **8.3** | **0** | **0.0** | | **8** | **53.3** | **5** | **41.7** | **3** | **100.0** | | **17** | **47.2** | **12** | **40.0** | **5** | **83.3** |
| ≤15 minute underestimate | 1 | 8.3 | 1 | 8.3 | 0 | 0.0 | | 5 | 33.3 | 2 | 16.7 | 3 | 100.0 | | 12 | 33.3 | 9 | 30.0 | 3 | 50.0 |
| ≤15 minute overestimate | 0 | 0.0 | 0 | 0.0 | 0 | 0.0 | | 3 | 20.0 | 3 | 25.0 | 0 | 0.0 | | 5 | 13.9 | 3 | 10.0 | 2 | 33.3 |
| ***Overestimates*** | **9** | **75** | **9** | **75** | **0** | **0.0** | | **6** | **40.0** | **6** | **50.0** | **0** | **0.0** | | **15** | **41.7** | **15** | **50.0** | **0** | **0.0** |
| ≥60 minute overestimate | 7 | 58.3 | 7 | 58.3 | 0 | 0.0 | | 1 | 6.7 | 1 | 8.3 | 0 | 0.0 | | 8 | 22.2 | 8 | 26.7 | 0 | 0.0 |
| 30-59 minute overestimate | 2 | 16.7 | 2 | 16.7 | 0 | 0.0 | | 1 | 6.7 | 1 | 8.3 | 0 | 0.0 | | 3 | 8.3 | 3 | 10.0 | 0 | 0.0 |
| 16-29 minute overestimate | 0 | 0.0 | 0 | 0.0 | 0 | 0.0 | | 4 | 26.7 | 4 | 33.3 | 0 | 0.0 | | 4 | 11.1 | 4 | 13.3 | 0 | 0.0 |
| **Discordance** |  | |  | |  | |  | |  | |  | |  | |  | |  | |  | |
| ***No time discordance*** | **11** | **91.7** | **11** | **91.7** | **0** | **0.0** | | **11** | **73.3** | **11** | **91.7** | **0** | **0.0** | | **24** | **66.7** | **24** | **80.0** | **0** | **0.0** |
| Estimated and actual both ≤ 30min | 0 | 0.0 | 0 | 0.0 | 0 | 0.0 | | 1 | 6.7 | 1 | 8.3 | 0 | 0.0 | | 3 | 8.3 | 3 | 10.0 | 0 | 0.0 |
| Estimated and actual both > 30min | 11 | 91.7 | 11 | 91.7 | 0 | 0.0 | | 10 | 66.7 | 10 | 83.3 | 0 | 0.0 | | 21 | 58.3 | 21 | 70.0 | 0 | 0.0 |
| ***Time discordance*** | **1** | **8.3** | **1** | **8.3** | **0** | **0.0** | | **4** | **26.7** | **1** | **8.3** | **3** | **100.0** | | **12** | **33.33** | **6** | **20.0** | **6** | **100.0** |
| Estimate at or under 30min, actual over 30min | 0 | 0.0 | 0 | 0.0 | 0 | 0.0 | | 1 | 6.7 | 1 | 8.3 | 0 | 0.0 | | 5 | 13.9 | 5 | 16.7 | 0 | 0.0 |
| Estimate over 30min, actual at or under 30min | 1 | 8.3 | 1 | 8.3 | 0 | 0.0 | | 3 | 20.0 | 0 | 0.0 | 3 | 100.0 | | 7 | 19.4 | 1 | 3.3 | 6 | 100.0 |
| ^1^ Data from 10 women were excluded because they did not collect water to bring home (9 Guatemala, 1 Kenya) and from 10 women because they did not provide estimated times (5 Guatemala, 1 Honduras, 4 Zimbabwe). Data from 38 women were excluded because their water source type was not the same for their estimated and measured water journey times (5 Guatemala, 10 Honduras, 9 Kenya, 14 Zimbabwe). For the Guatemala participant who completed two water journeys, only her water journey to collect water was included. | | | | | | | | | | | | | | | | | | | | |
